# Anti-PF4 VITT antibodies are oligoclonal and variably inhibited by heparin

**DOI:** 10.1101/2021.09.23.21263047

**Authors:** B Singh, A Kanack, A Bayas, G George, MY Abou-Ismail, M Kohlhagen, M Christ, M Naumann, K Moser, K Smock, A Grazioli, D Murray, A Padmanabhan

## Abstract

**Background:** COVID-19 vaccines have been associated with a rare thrombotic and thrombocytopenic reaction, Vaccine-induced immune thrombotic thrombocytopenia (VITT) characterized by platelet-activating anti-PF4 antibodies. This study sought to assess clonality of VITT antibodies and evaluate their characteristics in antigen-based and functional platelet studies.

**Methods:** Anti-PF4 antibodies were isolated from five patients with VITT secondary to ChAdOx1 nCoV-19 (n=1) or Ad26.COV2.S (n=4) vaccination. For comparative studies with heparin-induced thrombocytopenia (HIT), anti-PF4 antibodies were isolated from one patient with spontaneous HIT, another with “classical” HIT, and two patients with non-pathogenic (non-platelet activating) anti-PF4 antibodies. Isolated antibodies were subject to ELISA and functional testing, and mass spectrometric evaluation for clonality determination.

**Results:** All five VITT patients had oligoclonal anti-PF4 antibodies (3 monoclonal, one bi- and one tri-clonal antibodies), while HIT anti-PF4 antibodies were polyclonal. Notably, like VITT antibodies, anti-PF4 antibodies from a spontaneous HIT patient were monoclonal. The techniques employed did not detect non-pathogenic anti-PF4 antibodies. The ChAdOx1 nCoV-19-associated VITT patient made an excellent recovery with heparin treatment. In vitro studies demonstrated strong inhibition of VITT antibody-induced platelet activation with therapeutic concentrations of heparin in this and one Ad26.COV2.S-associated VITT patient. Oligoclonal VITT antibodies with persistent platelet-activating potential were detected at 6 and 10 weeks after acute presentation in two patients tested. Two of the 5 VITT patients had recurrence of thrombocytopenia and one patient had focal seizures several weeks after acute presentation.

**Conclusion:** Oligoclonal anti-PF4 antibodies mediate VITT. Heparin use in VITT needs to be further studied.

## Introduction

COVID-19 has caused significant challenges to health worldwide and has exacted a high toll on morbidity and mortality^1^. Several vaccines have shown high efficacy in clinical studies and have produced salutary effects on hospitalizations and deaths^2^. Two vaccines, ChAdOx1 nCoV-19 (AstraZeneca) and Ad26.COV2.S (Janssen/Johnson & Johnson) have been associated with very rare adverse reactions variably called Vaccine-induced immune thrombotic thrombocytopenia (VITT) and Thrombosis and Thrombocytopenia syndrome (TTS), respectively^3-20^. In this report, both syndromes are referred to as VITT, in line with expert recommendations^18^. VITT is characterized by strong anti-PF4 antibodies, thrombocytopenia, and thrombosis in almost all patients. Mortality rates are high at 22% in the largest study to date^21^, and even higher in others^3-5^. While VITT shares a key feature with another well-studied entity, Heparin-induced Thrombocytopenia (HIT)^22^ in that the generated antibodies recognize PF4, some differences have been noted. VITT antibodies are characterized by extremely high antibody optical densities in solid-phase PF4/Polyanion enzyme-linked immunosorbent assays (ELISAs), and interestingly, many antibodies do not activate heparin-treated platelets but rather require PF4-treated platelets for consistent detection in functional testing^3,15,20,23^. Clinically, thrombosis occurs in ∼20-60% of patients with HIT,^24^ while platelet-activating anti-PF4 antibodies in the vaccine setting almost always cause thrombosis, with only rare exceptions noted to date^5,10^.

Given this unique set of laboratory and clinical features, we embarked on a study to characterize pathogenic anti-PF4 antibodies in VITT patients. Results presented in this report show that in contrast to HIT where polyclonal anti-PF4 antibodies are seen, VITT patients produce monoclonal/oligoclonal anti-PF4 antibodies.

## Methods

### Patient Samples

Blood samples were obtained from patients suspected of thrombotic thrombocytopenia after vaccination with ChAdOx1 nCoV-19 (one patient), Ad26.COV2.S (four patients) and from patients with “classical” HIT (HIT that developed after heparin exposure), spontaneous (autoimmune) HIT and patients with non-pathogenic anti-PF4 antibodies. Samples were drawn during acute presentation from Patients 2-5, and ∼10 weeks post-acute presentation from Patient 1. A follow up sample from Patient 4, ∼6 weeks after acute presentation was also obtained. HIT patient samples were obtained at the time of acute presentation. Participants consented to publication of the research work. Research studies were approved by the Institutional Review Board of Mayo Clinic, Rochester, MN.

### Anti-PF4 antibody isolation

Heparin Sepharose beads (200mcL, Cytiva Lifesciences) were thoroughly washed with phosphate buffered saline, pH 7.4 (PBS) and incubated with 200µg of recombinant PF4 (Protein Foundry) or an equal volume of PBS (control condition). After incubation for 1hr, beads were incubated further with 0.5mL of patient sample (serum/plasma) for 1hour. Beads were then thoroughly washed with PBS and elution from the beads was performed with 0.5 mL 2M NaCl. The eluate was dialyzed against PBS before being evaluated by ELISA, PF4-dependent P-selectin Expression Assay (PEA) and mass spectrometric studies as noted below.

### ELISA and Functional platelet studies

ELISA plates (Thermo Scientific) were incubated with recombinant PF4 (Protein Foundry; 10 µg/mL) alone, PF4 and Polyvinyl sulfonate (PVS, Polysciences; 9 µg/mL) or PF4 and Unfractionated heparin (Sigma Aldrich; 0.4U/mL). Plates were washed with PBS 0.1% Tween and blocked with Superblock T20 (Thermo Scientific). Eluate samples were tested at a 1:10 dilution while serum/plasma samples were tested at the standard 1:50 dilution. Goat anti-human IgG fc antibody (Jackson Immunoresearch) and pNPP (Sigma Aldrich) were used for colorimetric detection. Optical density was recorded at 405 nm. The PF4-dependent p-selectin expression assay (PEA) was performed as previously described^25^.

### Liquid Chromatography Electrospray Ionization Quadrupole time-of-flight mass spectrometry (LC-ESI-QTOF MS)

This technique was performed as described elsewhere^26-28^. Briefly, Immunoglobulins (Igs) from patient serum/plasma or eluates from PF4-Heparin beads/Control Heparin beads were first immunoenriched using camelid-derived nanobodies directed towards the constant region of IgG heavy chain, kappa, or lambda light chains, reduced, and subjected to liquid chromatography for separation of immunoglobulins prior to ESI-QTOF-MS. A detailed description of the methods is included in the *Supplemental Methods* section.

### Immunofixation Electrophoresis

Serum IFE was performed using Hydrasys 9IF gels (Sebia, Paris, France) following manufacturer’s recommendations.

## Results

### VITT patient presentation and course

*Supplementary Appendix* and **Fig S1** provide details of the 5 VITT patients described in this report. Briefly, all five patients presented with thrombocytopenia and thrombosis. All except Patient 1 were treated with IVIg. Patient 1 was treated with dexamethasone and unfractionated heparin, followed by enoxaparin during bridging to phenprocoumon which was associated with a steady and sustained increase in platelet count. All patients received corticosteroids. Patients 2-5 also received parenteral direct thrombin inhibitors briefly followed by direct oral anticoagulants. While IVIg was effective in increasing platelet counts in the four treated patients, two patients, Patient 2 and 4 had recurrence of thrombocytopenia. Redosing of IVIg was ineffective when it was attempted in Patient 2, and thrombocytopenia was protracted in this patient requiring >2 months for platelet counts to reach 150,000× 10^9^/L. Patient 1 experienced focal seizures ∼3 months after initial presentation and was noted to have CVST, not identified on initial admission. All patients had strong positive results in solid phase ELISA, but serotonin release assay (SRA) results were inconsistent (**Fig S1**).

### Antibody binding to PF4 antigen targets and platelet activation

Patient samples were tested in an ELISA format to evaluate if VITT antibodies recognize uncomplexed PF4 and PF4 complexed with the polyanions, polyvinyl sulfonate, and heparin, the latter forming the basis of currently used HIT diagnostic assays. In contrast to HIT antibodies, antibodies from VITT patients recognized uncomplexed PF4, in addition to recognizing complexes of PF4 with polyanions (**Fig 1A**). In fact, the optical density was highest with uncomplexed PF4 relative to complexed PF4 across all VITT patients. The PEA demonstrated VITT sample mediated platelet activation in all five patients, including Patient 1 whose sample was drawn >2 months after initial presentation (**Fig 1B**).

**Figure 1.**
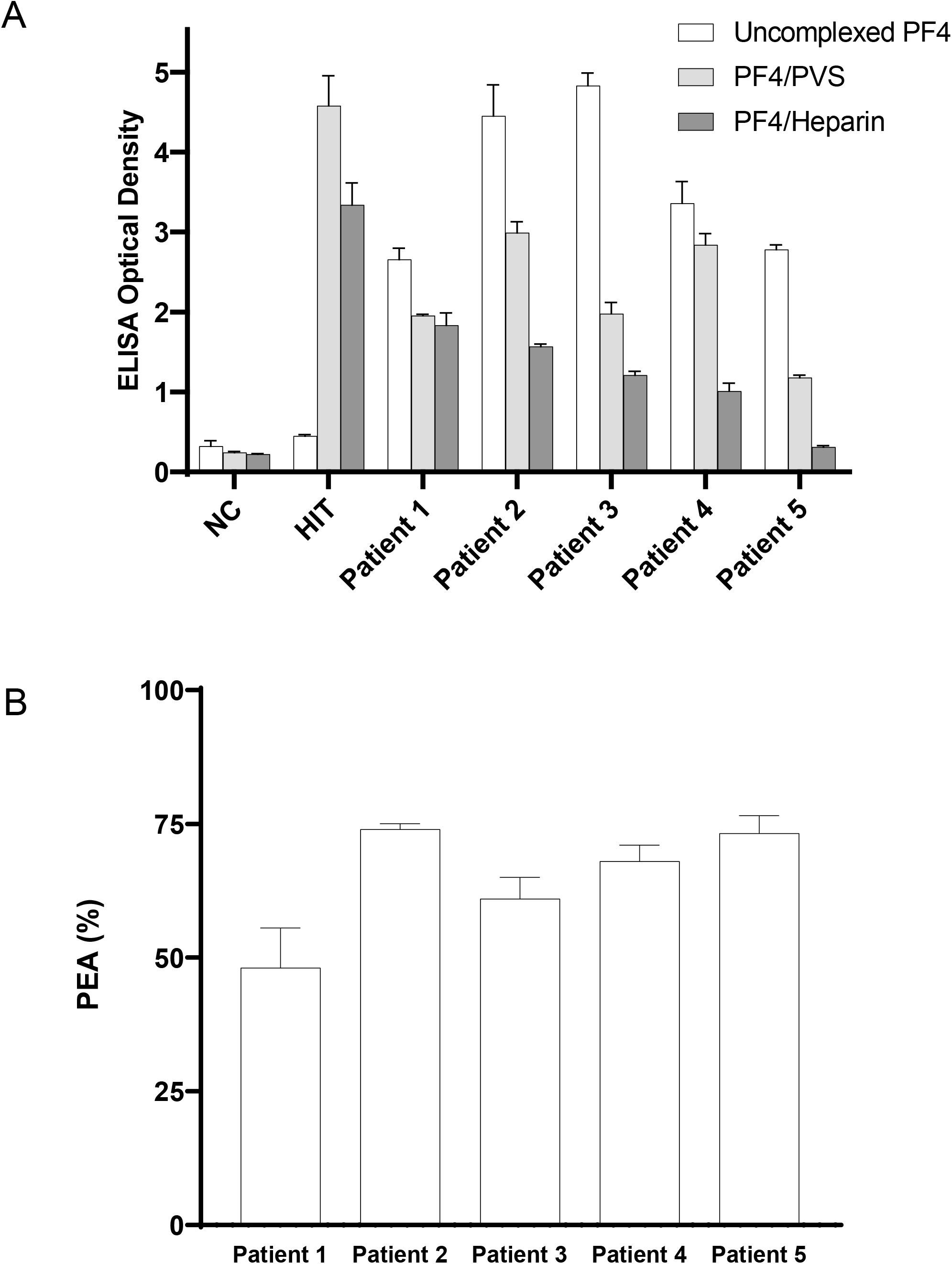
VITT antibodies recognize uncomplexed PF4 and activate PF4-treated platelets. **(A)** Antibody binding to PF4 alone (“uncomplexed”; white), PF4/Polyvinylsulfonate (light gray), or PF4/Unfractionated heparin (dark gray) targets were evaluated by ELISA. **(B)** Activation of VITT patient antibodies in the PF4-dependent P-selectin Expression Assay (PEA) was examined. Normal Control (NC), HIT Patient (HIT). Means and SD (n=3) are presented.

### Isolated anti-PF4 antibodies are platelet activating and are non-uniformly inhibited by heparin

Techniques used in these studies are schematically presented in **Fig 2A-B** and described in the *Methods* section. VITT anti-PF4 antibodies eluted from PF4-heparin or heparin (control) beads were first validated in the PF4/polyanion ELISA (**Fig 3A**) and PEA (**Fig 3B**). Results demonstrated that antibodies eluted from PF4-heparin treated beads bound PF4/polyanion complexes strongly and activated platelets in the PEA. No anti-PF4 antibodies/platelet-activating antibodies were noted in eluates from heparin-sepharose beads not treated with PF4 (**Fig 3A-B**). Given the excellent platelet recovery during heparin therapy in Patient 1 without the need for IVIg, we evaluated the ability of therapeutic concentrations of heparin to inhibit VITT anti-PF4 ab mediated platelet activation (**Fig 3C**). Notably, heparin inhibited platelet activation induced by Patient 1 and Patient 5 but not Patients 2-4, at therapeutic concentrations.

**Figure 2.**
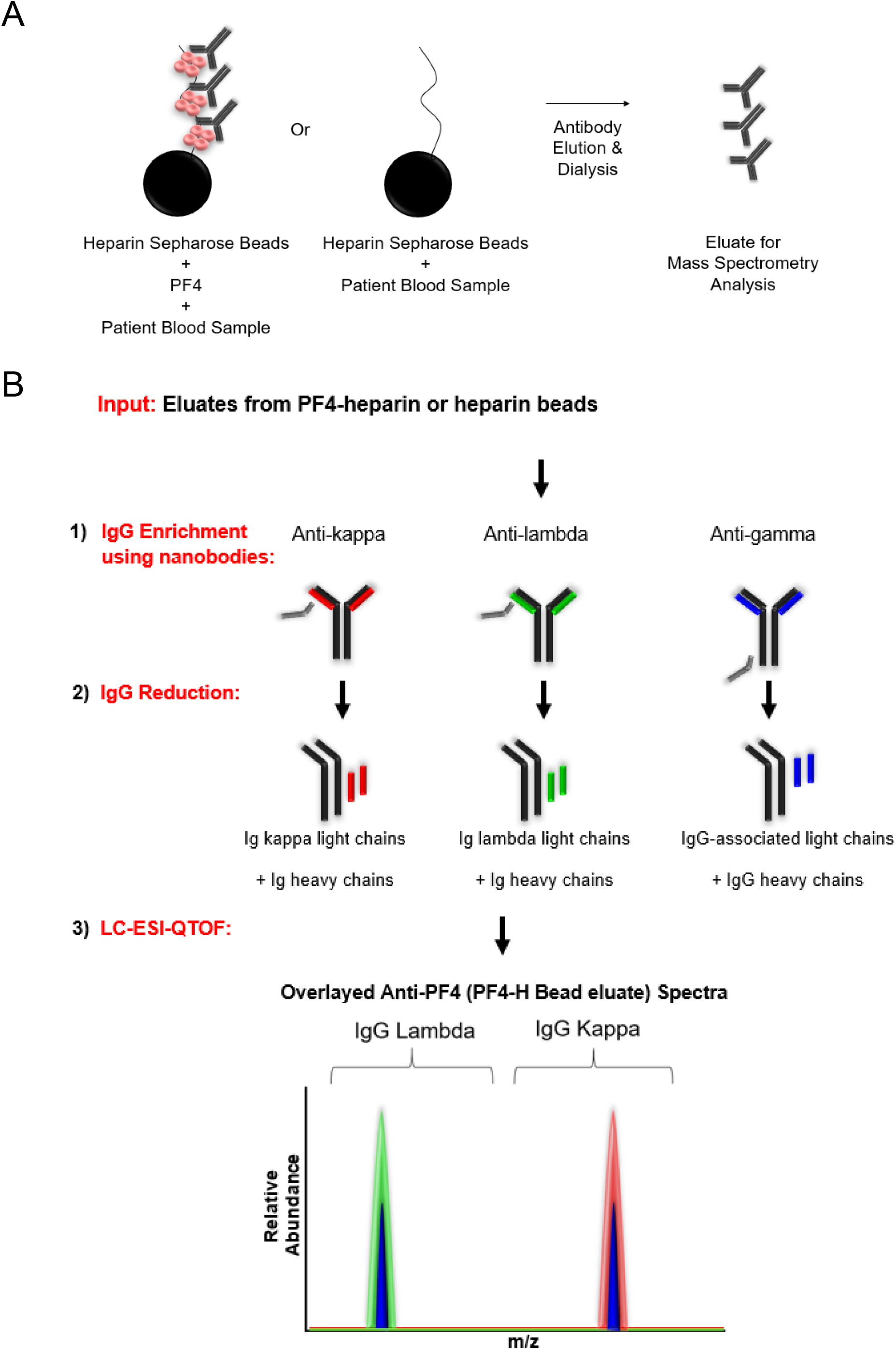
Strategy for anti-PF4 antibody isolation and clonality assessment. **(A)** Patient sera were incubated with PF4-heparin sepharose beads or heparin (control) sepharose beads. Antibodies were eluted by high salt concentration and dialyzed. This eluate was subject to mass spectrometric analysis. **(B)** Immunoglobulins (Igs) eluted from PF4-heparin sepharose or heparin sepharose beads were (1) Isolated using camelid nanobodies (gray) specific for kappa light chains (red), lambda light chains (green), or gamma heavy chains (black). IgG-associated light chains immunoenriched with anti-gamma heavy chain antibodies are shown in blue (right). Isolated Igs were then (2) reduced and (3) analyzed using liquid chromatography-electrospray ionization-quadrupole time-of-flight mass spectrometry (LC-ESI-QTOF) to determine the antibody repertoire present. In the spectra, green represents the light chain mass to charge (m/z) distribution of all lambda containing Igs, red represents the light chain (m/z) distribution of all kappa containing Igs, and blue represents the light chain (m/z) distribution of kappa and lambda light chains associated with an IgG heavy chain. Spectra are overlaid to confirm the type of light chain and antibody isotype. In this example, the anti-PF4 antibody is biclonal, with one IgG lambda and one IgG kappa monoclonal antibody.

**Figure 3.**
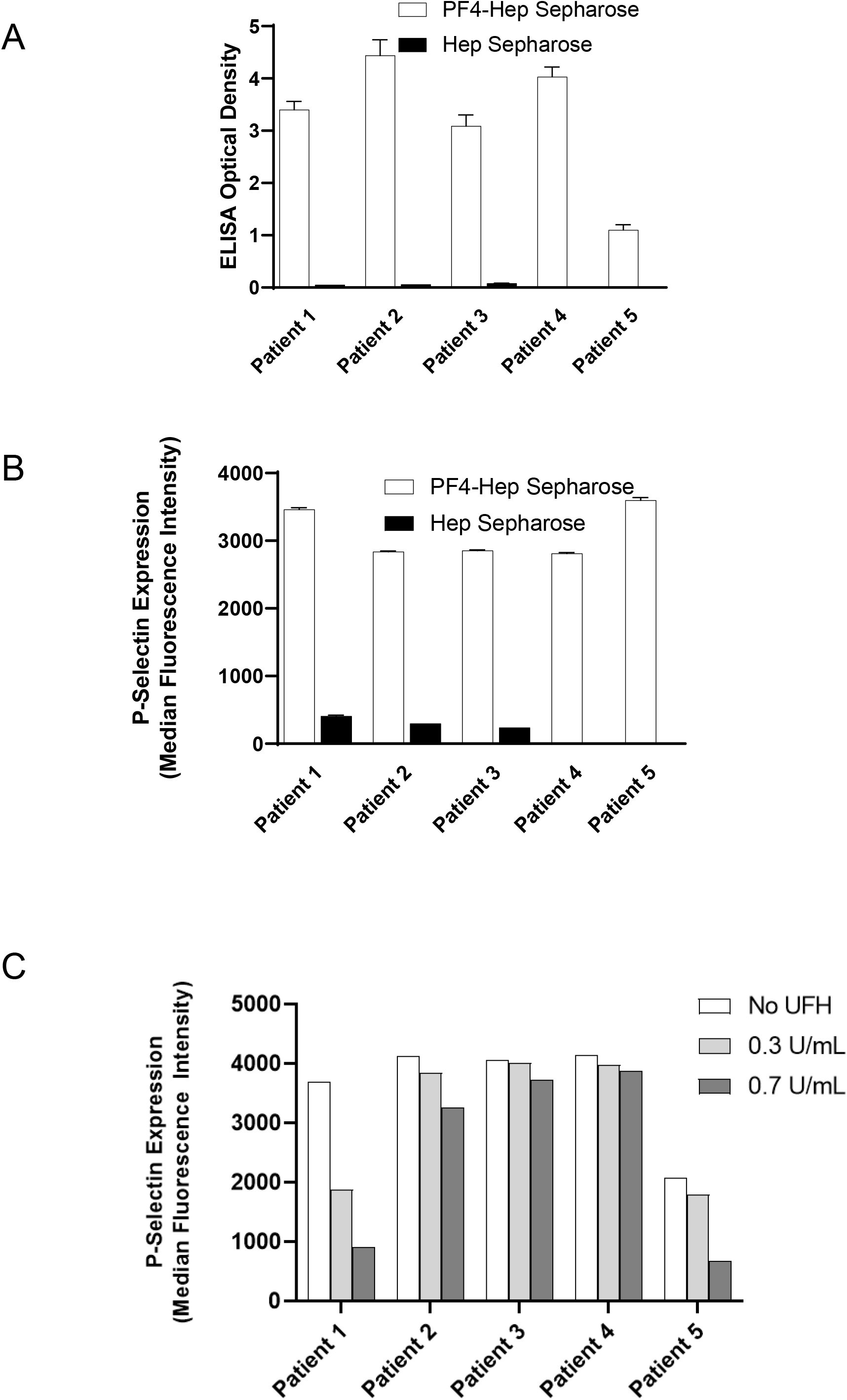
Eluted antibodies recognize PF4, activate platelets and are variably heparin-inhibitable. Eluates from PF4-heparin beads and control heparin beads were evaluated in the ELISA plated with PF4/Polyvinylsulfonate complexes (**A**) and for platelet activation in the PEA (**B**). Impact of unfractionated heparin (UFH) at 0.3U/mL and 0.7U/mL on VITT anti-PF4 antibody mediated platelet activation is shown in (**C**). Means and SD (n=3) are presented in (**A**) and (**B**), while results from a single experiment are presented in (**C**). Control (heparin) bead studies were not performed with Patients 4 and 5 (in **A** and **B**) due to limited sample volume.

### VITT anti-PF4 antibody clonality assessment

LC-ESI-QTOF mass spectrometry was performed on the eluates from PF4-heparin and control beads. Results demonstrated monoclonal anti-PF4 antibodies in Patients 1, 2 and 5 (**Fig 4A-B and 4E**), while biclonal and triclonal anti-PF4 antibodies were noted in Patients 3 and 4, respectively (**Fig 4B-C)**. Of note, the monoclonal antibody seen in Patient 1 was lower in abundance compared to the other VITT patients relative to background which was not surprising as the sample was drawn ∼10 weeks after acute presentation (**Fig 4A**). Anti-PF4 antibodies from all VITT patients had the *lambda* light chain. These oligoclonal antibodies, while obvious upon evaluation of the isolated anti-PF4 antibody, were not detectable upon evaluation of whole serum IgG from the patient: Immunofixation electrophoresis, performed on samples from patients 1 and 2, did not identify monoclonal or oligoclonal bands (**Fig S2**). Similarly, **Fig S3A** shows that the monoclonal antibody from Patient 2 is barely appreciable above the polyclonal background.

**Figure 4.**
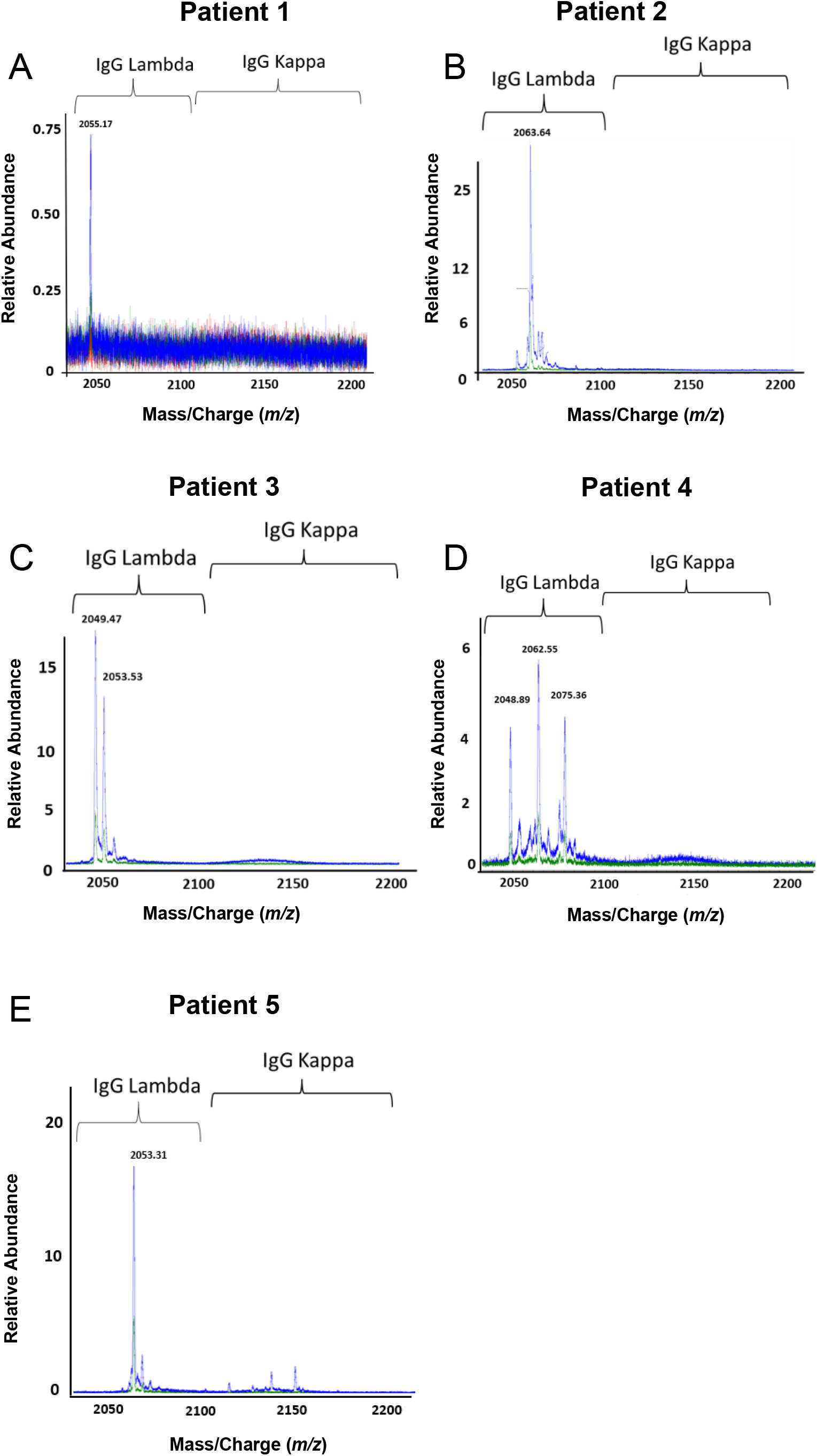
VITT patients produce oligoclonal anti-PF4 antibodies. Displayed are LC-ESI-QTOF MS light chain +11 (m/z, mass to charge) distributions from anti-PF4 antibodies isolated from five patients with VITT. In the spectra, green represents the distribution of all lambda containing immunoglobulins (Igs), red represents the +11 m/z distribution of all kappa containing Igs, and blue represents the +11 m/z light chain distribution of kappa and lambda light chains associated with an IgG heavy chain. The number listed above peaks indicate the +11 mass/charge (m/z) ratio of the identified light chain. The X-axis shows mass/charge ratios and Y-axis depicts the relative abundance of the monoclonal/oligoclonal antibody identified.

### Antibody clonality in Spontaneous HIT, Classical HIT and Patients with non-pathogenic anti-PF4 antibodies

Studies were performed to assess antibody clonality in a patient who developed spontaneous HIT (autoimmune, aHIT with ELISA OD of 2.782 and SRA-positive)^29^ which, like VITT, develops in the absence of proximate heparin treatment, as well as a patient with classical *heparin-induced* HIT (ELISA OD of 2.500 and SRA-positive) and two patients who did not have HIT but developed non-pathogenic antibodies after heparin exposure during cardiac surgery (Non-pathogenic HIT, NP-HIT; ELISA-positive with OD of 0.429 [NP-HIT1] and 0.426 [NP-HIT2], both SRA-negative). First, Anti-PF4 antibody isolation was performed as described above and isolated antibodies were tested in PEA and ELISA. HIT and aHIT antibodies strongly activated platelets, as expected in the PEA (**Fig 5A**. For reference MFI from a strong platelet agonist, Thrombin receptor activating peptide, TRAP was ∼4000). As expected, bead eluates from the two patients with non-pathogenic anti-PF4 antibodies did not activate platelets (**Fig 5A**). Testing in the PF4/polyanion ELISA demonstrated strong binding of HIT and aHIT antibodies, and modest binding of NP-HIT antibodies in PF4-heparin bead eluates vs heparin (control) bead eluates suggesting that anti-PF4 antibodies were present in all eluates tested (**Fig 5B**). Mass spectrometric evaluation of PF4-heparin bead eluate from the aHIT patient demonstrated a strong IgG kappa monoclonal anti-PF4 antibody (**Fig 5C**). Consistent with current dogma, the HIT patient’s anti-PF4 antibodies were polyclonal (**Fig 5D**), despite which very strong platelet activation was seen (**Fig 5A**). Anti-PF4 antibodies from the two patients with non-activating anti-PF4 antibodies were below level of detection in mass spectrometry (**Fig 5E-F**). Of note, evaluation of the entire serum IgG repertoire of one of the patients with non-pathogenic HIT antibodies demonstrated a monoclonal antibody (**Fig S3B**) that was not pulled down non-specifically by our anti-PF4 isolation techniques. In addition, eluates from control (heparin sepharose) beads evaluated by mass spectrometry showed no eluted immunoglobulins, showing that PF4 bound to beads was critical for isolation of anti-PF4 antibodies (**Fig S4A-D**). **VITT antibodies are persistent** Patient 4 provided follow-up samples ∼6 weeks after initial presentation. Testing revealed persistent antibodies that recognized uncomplexed PF4 and PF4-polyanion complexes, although at lower levels compared to the acute samples (compare **Fig S5A** to **Fig 1A**).

**Figure 5.**
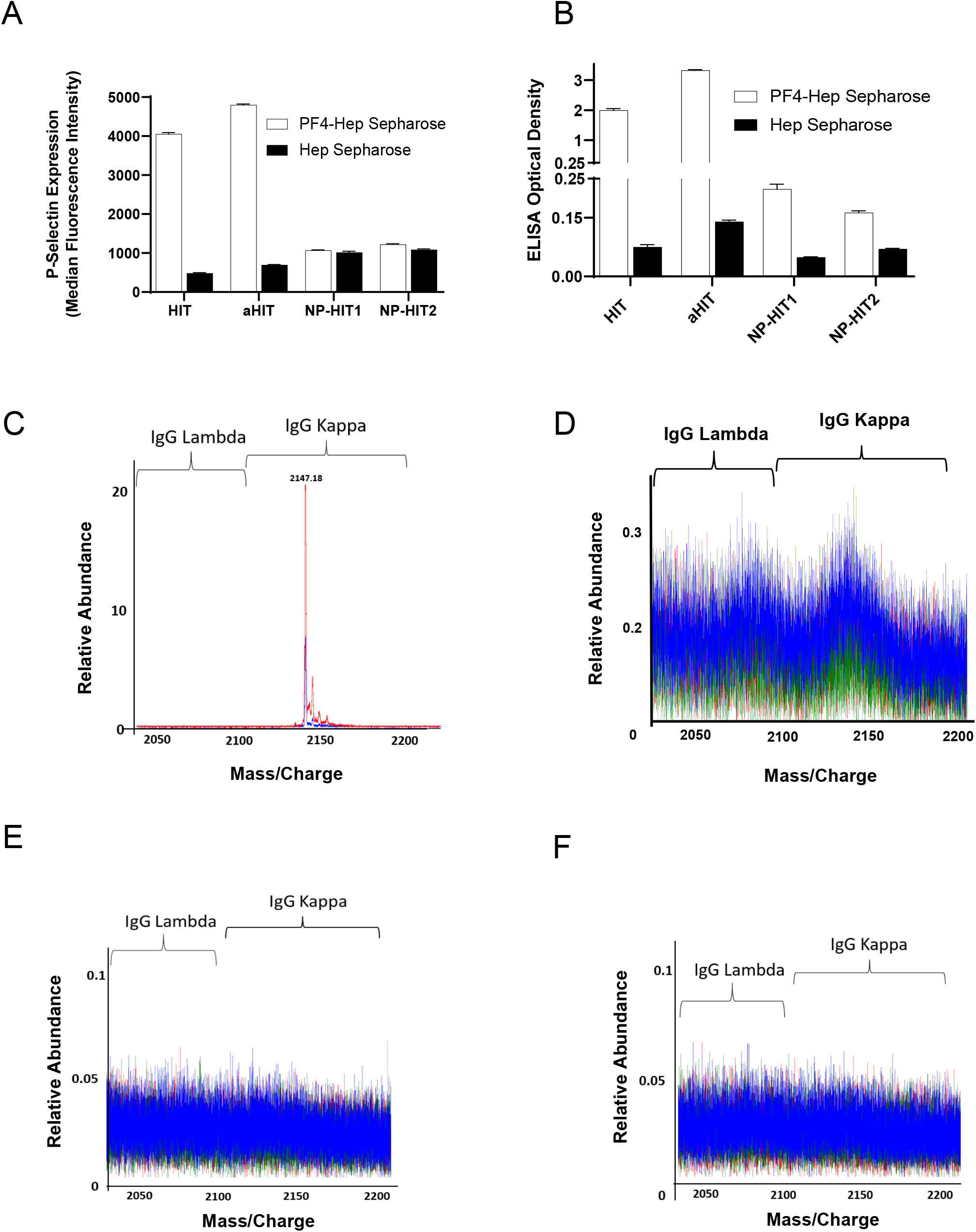
Anti-PF4 antibody characterization in aHIT, HIT and patients with non-pathogenic (NP) HIT antibodies. Eluates from PF4-heparin beads and control heparin beads were evaluated for platelet activation in the PEA (**A**) and PF4/Polyanion ELISA (**B**). Means and SD (n=3) are presented in (**A**) and (**B**). **C-F**: Displayed are LC-ESI-QTOF MS +11 light chain distributions from anti-PF4 antibodies isolated from patients with aHIT (**C**), HIT (**D**) and NP-HIT (**E-F**). In the spectra, green represents the distribution of all lambda containing immunoglobulins (Igs), red represents the distribution of all kappa containing Igs, and blue represents the light chain distribution of kappa and lambda light chains associated with an IgG heavy chain. The number listed above peaks depicts the mass/charge (m/z) ratio of the identified light chain. The X-axis shows mass/charge ratios and Y-axis depicts the relative abundance of the monoclonal/oligoclonal antibody identified.

Similarly, while still platelet-activating, p-selectin expression was lower in the follow-up sample relative to the acute serum (compare **Fig S5B** to **Fig 1B**). Tri-clonal HIT antibodies were still detectable by mass spectrometry, but in lower abundance (compare Y axis in **Fig S5C** to Fig **4D**).

## Discussion

Murray *et al* have recently used mass spectrometry for the sensitive and specific detection of monoclonal proteins in multiple myeloma and related disorders.^30,31^ In this study, we isolated anti-PF4 antibodies using heparin-PF4 beads and coupled it to mass spectrometry to assess antibody clonality. To exclude the possibility that the techniques used to isolate anti-PF4 antibodies resulted in the selective enrichment of only those antibodies that had the highest affinity for PF4 that were then visualized as oligoclonal antibodies by mass spectrometry, we tested samples from a patient with classical HIT where current dogma suggest polyclonality of anti-PF4 antibodies^32-34^. Isolated anti-PF4 antibodies from HIT strongly activated platelets and were confirmed to be polyclonal. A sensitive technique such as mass spectrometry may potentially detect irrelevant antibodies such as those seen in patients with PF4/polyanion ELISA-positive, SRA-negative anti-PF4 antibodies. Thus, we also evaluated samples from two patients who developed non-pathogenic anti-PF4 antibodies after heparin exposure. Results demonstrated that anti-PF4 antibodies from these patients were below the limit of detection, suggesting high specificity of the techniques used. Therefore, the finding of mono- and oligoclonal antibodies in VITT and spontaneous HIT patients is likely not an artifact of the novel application of this technique in this field. Unlike chronic monoclonal gammopathies like multiple myeloma where antibody levels increase over months/years, VITT/spontaneous HIT is an acute event. Thus, it was not surprising that levels of monoclonal/oligoclonal antibodies were not high enough to be detected upon testing of the patient’s entire IgG repertoire: It was critical to couple anti-PF4 antibody isolation to downstream mass spectrometry for proper recognition of antibody clonality.

The observation of monoclonal/oligoclonal anti-PF4 antibodies in ChAdOx1 nCoV-19-associated and Ad26.COV2.S-associated VITT points to both disorders being similar. All five patients’ monoclonal/oligoclonal antibodies had exclusively lambda light chains. Based on the limited number of samples, it is too early to know if lambda light chain restriction is characteristic of this syndrome, and it is unclear how this may impact disease pathophysiology. Like VITT, the spontaneous HIT patient who developed thrombosis and thrombocytopenia after an infectious prodrome in the absence of heparin exposure and was found to have monoclonal anti-PF4 antibodies (although with kappa light chain). Recent research suggests that healthy subjects possess preexisting inactive PF4/heparin-specific B cells,^35^ and it is conceivable that single/few preexisting anti-PF4 B cell clones are activated after proinflammatory stimuli such as vaccination, orthopedic surgery, or infections causing VITT/spontaneous HIT. These hypotheses should be addressed in future studies.

The natural history of VITT antibodies is not yet known. While most VITT patients tested in this study had samples obtained during their acute episode, studies were performed on Patients 1 and 4 ten and six weeks after acute presentation, respectively. Results revealed persistent platelet activating and strongly ELISA binding antibodies in both native sera and isolated anti-PF4 antibody fractions in both patients. Patient 4’s recovery was complicated by a recurrence of thrombocytopenia, and Patient 1 had focal seizures ∼3 months after acute presentation. A third patient, Patient 2 also had recurrence of thrombocytopenia with a nadir of 101 × 10^9^/L ∼30 days after acute presentation which persisted at <150,000 × 10^9^/L for another ∼70 days despite IVIg re-dosing.

Recent studies demonstrate that several VITT patients experience refractoriness to maximal therapy (including IVIg) or relapse requiring additional treatments such as therapeutic plasma exchange (TPE) and rituximab^21,36^. Of note, 17 patients (8% of those classified as having definite/probable VITT) in the most extensive VITT study to date required TPE, which was associated with a 90% survival rate suggesting a salutary effect of this intervention^21^. In fact, the recently released National Institute for Health and Care excellence guideline on VITT notes that relapses after initial response to treatment can happen quickly and makes recommendations for monthly monitoring for at least 6 months after which monitoring can be performed quarterly if no relapses were detected^37^. Persistent oligoclonal anti-PF4 antibodies that develop in VITT, as shown in this study, may bind strongly to PF4 targets^38^, mediate severe thrombosis and thrombocytopenia and cause refractoriness to IVIg. Together, these factors may explain the high mortality of >20% seen in VITT patients^21^, relative to ∼10% seen in HIT^39^.

Therapeutic concentrations of heparin were able to significantly inhibit anti-PF4-mediated platelet activation in two of our five VITT patients. Of note, one of these two patients developed VITT after ChAdOx1 nCoV-19 vaccination and received heparin at acute presentation for ∼two weeks following by enoxaparin for an additional week and had a prompt and durable recovery of platelet counts from a presenting count of 30 × 10^9^/L. A role for dexamethasone in mediating this recovery was deemed unlikely due to the very short period of administration (4 days). This data on inhibition of VITT antibodies by heparin is consistent with a recent report that demonstrates that ChAdOx1 nCoV-19-associated VITT antibodies bind PF4 within its heparin-binding site^38^.

However, anti-PF4 antibodies from only one of four Ad26.COV2.S-associated patients were inhibited by heparin in functional testing suggesting that anti-PF4 antibodies in these patients may recognize additional PF4 epitopes, outside the heparin binding domain. Current clinical guidance recommends against use of heparin in all patients with VITT^40^. Our data suggest that this guidance may need to be reassessed. In fact, in a recent report^21^, 23% of patients (n=50) with definite/probable VITT received heparin which did not appear to be deleterious, although numbers were too small to draw conclusions on efficacy. Additional case reports appear to confirm the lack of adverse effect of heparin therapy in VITT patients^11^. Thus, heparin could be an additional therapeutic modality particularly in severely afflicted patients refractory to currently used therapies. We also demonstrate aberrant characteristics of VITT antibodies relative to those seen in HIT. First, VITT antibodies have a very high level of binding to PF4-Polyanion complexes as evidenced by extremely high ELISA optical densities. Second, they bind uncomplexed PF4, and third, VITT antibodies only sometimes activate heparin-treated platelets (as in the SRA), but consistently activate PF4-treated platelets. This body of data is consistent with those recently reported by other groups^3,15,20,23^.

In summary, this study demonstrates that VITT patients produce strong, persistent platelet-activating mono- and oligoclonal anti-PF4 antibodies and suggests that long-term follow up of these patients may be indicated. Efforts towards early identification of VITT patients who may require more aggressive treatment should be undertaken, and the role of heparin treatment in this syndrome should be investigated.

## Data Availability

Data will be made available upon reasonable request.

## Acknowledgments

We would like to thank Stephanie Hafner, BS from Mayo Clinic’s Research Innovation Office for exceptional research coordination support. This work was supported, in part, by National Institutes of Health grant HL133479 (A.P.).

## Authorship

BS, AK, AB, GG, YAI and AG provided critical input on laboratory and clinical elements of the study. BS and AK performed the studies described on anti-PF4 antibody isolation, ELISA studies and assessment of platelet activation. MK performed mass spectrometry studies. MC, MN, KM and KS provided helpful advice on clinical and laboratory aspects of the manuscript. DM and AP conceived the experimental plan and directed the laboratory studies. BS, AK, and AP wrote the first draft and all authors provided input and approved the final version.

## Conflict-of-interest disclosure

AP reports pending/issued patents (Mayo Clinic, Retham Technologies and Versiti BloodCenter of Wisconsin), equity ownership in Retham Technologies, and serving on the advisory board of Veralox Therapeutics. DM reports pending/issued patents (Mayo Clinic). The remaining authors declare no competing financial interests.

## Notes

### Author Declarations

Mayo Clinic Institutional Review Board

## References

1. Wiersinga WJ, Rhodes A, Cheng AC, Peacock SJ, Prescott HC. Pathophysiology, Transmission, Diagnosis, and Treatment of Coronavirus Disease 2019 (COVID-19): A Review. JAMA 2020;324(8):782–793. DOI: 10.1001/jama.2020.12839.

2. Creech CB, Walker SC, Samuels RJ. SARS-CoV-2 Vaccines. JAMA 2021;325(13):1318–1320. DOI: 10.1001/jama.2021.3199.

3. Greinacher A, Thiele T, Warkentin TE, Weisser K, Kyrle PA, Eichinger S. Thrombotic Thrombocytopenia after ChAdOx1 nCov-19 Vaccination. N Engl J Med 2021;384(22):2092–2101. DOI: 10.1056/NEJMoa2104840.

4. Schultz NH, Sorvoll IH, Michelsen AE, et al. Thrombosis and Thrombocytopenia after ChAdOx1 nCoV-19 Vaccination. N Engl J Med 2021;384(22):2124–2130. DOI: 10.1056/NEJMoa2104882.

5. Scully M, Singh D, Lown R, et al. Pathologic Antibodies to Platelet Factor 4 after ChAdOx1 nCoV-19 Vaccination. N Engl J Med 2021;384(23):2202–2211. DOI: 10.1056/NEJMoa2105385.

6. Tolboll Sorensen AL, Rolland M, Hartmann J, et al. A case of thrombocytopenia and multiple thromboses after vaccination with ChAdOx1 nCoV-19 against SARS-CoV-2. Blood Adv 2021;5(12):2569–2574. DOI: 10.1182/bloodadvances.2021004904.

7. Costello A, Pandita A, Devitt J. Case Report: Thrombotic Thrombocytopenia after COVID-19 Janssen Vaccination. Am Fam Physician 2021;103(11):646–647.

8. Muir KL, Kallam A, Koepsell SA, Gundabolu K. Thrombotic Thrombocytopenia after Ad26.COV2.S Vaccination. N Engl J Med 2021;384(20):1964–1965. DOI: 10.1056/NEJMc2105869.

9. See I, Su JR, Lale A, et al. US Case Reports of Cerebral Venous Sinus Thrombosis With Thrombocytopenia After Ad26.COV2.S Vaccination, March 2 to April 21, 2021. JAMA 2021;325(24):2448–2456. DOI: 10.1001/jama.2021.7517.

10. Thaler J, Ay C, Gleixner KV, et al. Successful treatment of vaccine-induced prothrombotic immune thrombocytopenia (VIPIT). J Thromb Haemost 2021. DOI: 10.1111/jth.15346.

11. Tiede A, Sachs UJ, Czwalinna A, et al. Prothrombotic immune thrombocytopenia after COVID-19 vaccine. Blood 2021. DOI: 10.1182/blood.2021011958.

12. Makris M, Pavord S, Lester W, Scully M, Hunt B. Vaccine-induced Immune Thrombocytopenia and Thrombosis (VITT). Res Pract Thromb Haemost 2021;5(5):e12529. DOI: 10.1002/rth2.12529.

13. Blauenfeldt RA, Kristensen SR, Ernstsen SL, Kristensen CCH, Simonsen CZ, Hvas AM. Thrombocytopenia with acute ischemic stroke and bleeding in a patient newly vaccinated with an adenoviral vector-based COVID-19 vaccine. J Thromb Haemost 2021. DOI: 10.1111/jth.15347.

14. Abou-Ismail MY, Moser KA, Smock KJ, Lim MY. Vaccine-induced thrombotic thrombocytopenia following Ad26.COV2.S vaccine in a man presenting as acute venous thromboembolism. Am J Hematol 2021. DOI: 10.1002/ajh.26265.

15. George G, Friedman KD, Curtis BR, Lind SE. Successful treatment of thrombotic thrombocytopenia with cerebral sinus venous thrombosis following Ad26.COV2.S vaccination. Am J Hematol 2021. DOI: 10.1002/ajh.26237.

16. Bayas A, Menacher M, Christ M, Behrens L, Rank A, Naumann M. Bilateral superior ophthalmic vein thrombosis, ischaemic stroke, and immune thrombocytopenia after ChAdOx1 nCoV-19 vaccination. Lancet 2021;397(10285):e11. DOI: 10.1016/S0140-6736(21)00872-2.

17. Muster V, Gary T, Raggam RB, Wolfler A, Brodmann M. Pulmonary embolism and thrombocytopenia following ChAdOx1 vaccination. Lancet 2021;397(10287):1842. DOI: 10.1016/S0140-6736(21)00871-0.

18. Arepally GM, Ortel TL. Vaccine-Induced Immune Thrombotic Thrombocytopenia (VITT): What We Know and Don’t Know. Blood 2021. DOI: 10.1182/blood.2021012152.

19. Cines DB, Bussel JB. SARS-CoV-2 Vaccine-Induced Immune Thrombotic Thrombocytopenia. N Engl J Med 2021;384(23):2254–2256. DOI: 10.1056/NEJMe2106315.

20. Bourguignon A, Arnold DM, Warkentin TE, et al. Adjunct Immune Globulin for Vaccine-Induced Thrombotic Thrombocytopenia. N Engl J Med 2021. DOI: 10.1056/NEJMoa2107051.

21. Pavord S, Scully M, Hunt BJ, et al. Clinical Features of Vaccine-Induced Immune Thrombocytopenia and Thrombosis. N Engl J Med 2021. DOI: 10.1056/NEJMoa2109908.

22. Warkentin TE. Laboratory diagnosis of heparin-induced thrombocytopenia. Int J Lab Hematol 2019;41 Suppl 1:15–25. DOI: 10.1111/ijlh.12993.

23. Vayne C, Rollin J, Gruel Y, et al. PF4 Immunoassays in Vaccine-Induced Thrombotic Thrombocytopenia. N Engl J Med 2021. DOI: 10.1056/NEJMc2106383.

24. Arepally GM, Padmanabhan A. Heparin-Induced Thrombocytopenia: A Focus on Thrombosis. Arterioscler Thromb Vasc Biol 2021;41(1):141–152. DOI: 10.1161/ATVBAHA.120.315445.

25. Samuelson Bannow B, Warad DM, Jones CG, et al. A prospective, blinded study of a PF4-dependent assay for HIT diagnosis. Blood 2021;137(8):1082–1089. DOI: 10.1182/blood.2020008195.

26. Kohlhagen M, Dasari S, Willrich M, et al. Automation and validation of a MALDI-TOF MS (Mass-Fix) replacement of immunofixation electrophoresis in the clinical lab. Clin Chem Lab Med 2020;59(1):155–163. DOI: 10.1515/cclm-2020-0581.

27. Barnidge DR, Dasari S, Ramirez-Alvarado M, et al. Phenotyping polyclonal kappa and lambda light chain molecular mass distributions in patient serum using mass spectrometry. J Proteome Res 2014;13(11):5198–205. DOI: 10.1021/pr5005967.

28. Barnidge DR, Dasari S, Botz CM, et al. Using mass spectrometry to monitor monoclonal immunoglobulins in patients with a monoclonal gammopathy. J Proteome Res 2014;13(3):1419–27. DOI: 10.1021/pr400985k.

29. Irani M, Siegal E, Jella A, Aster R, Padmanabhan A. Use of intravenous immunoglobulin G to treat spontaneous heparin-induced thrombocytopenia. Transfusion 2019;59(3):931–934. DOI: 10.1111/trf.15105.

30. Murray DL, Puig N, Kristinsson S, et al. Mass spectrometry for the evaluation of monoclonal proteins in multiple myeloma and related disorders: an International Myeloma Working Group Mass Spectrometry Committee Report. Blood Cancer J 2021;11(2):24. DOI: 10.1038/s41408-021-00408-4.

31. Murray DL, Dasari S. Clinical Mass Spectrometry Approaches to Myeloma and Amyloidosis. Clin Lab Med 2021;41(2):203–219. DOI: 10.1016/j.cll.2021.03.003.

32. Li ZQ, Liu W, Park KS, et al. Defining a second epitope for heparin-induced thrombocytopenia/thrombosis antibodies using KKO, a murine HIT-like monoclonal antibody. Blood 2002;99(4):1230–6.

33. Huynh A, Arnold DM, Kelton JG, et al. Characterization of platelet factor 4 amino acids that bind pathogenic antibodies in heparin-induced thrombocytopenia. J Thromb Haemost 2019;17(2):389–399. DOI: 10.1111/jth.14369.

34. Staibano P, Arnold DM, Bowdish DM, Nazy I. The unique immunological features of heparin-induced thrombocytopenia. Br J Haematol 2017;177(2):198–207. DOI: 10.1111/bjh.14603.

35. Zheng Y, Wang AW, Yu M, et al. B-cell tolerance regulates production of antibodies causing heparin-induced thrombocytopenia. Blood 2014;123(6):931–4. DOI: 10.1182/blood-2013-11-540781.

36. Patriquin CJ, Laroche V, Selby R, et al. Therapeutic Plasma Exchange in Vaccine-Induced Immune Thrombotic Thrombocytopenia. N Engl J Med 2021. DOI: 10.1056/NEJMc2109465.

37. https://www.nice.org.uk/guidance/ng200. Accessed 8/25/21.

38. Huynh A, Kelton JG, Arnold DM, Daka M, Nazy I. Antibody epitopes in vaccine-induced immune thrombotic thrombocytopaenia. Nature 2021. DOI: 10.1038/s41586-021-03744-4.

39. Dhakal B, Kreuziger LB, Rein L, et al. Disease burden, complication rates, and health-care costs of heparin-induced thrombocytopenia in the USA: a population-based study. Lancet Haematol 2018;5(5):e220–e231. DOI: 10.1016/S2352-3026(18)30046-2.

40. Oldenburg J, Klamroth R, Langer F, et al. Diagnosis and Management of Vaccine-Related Thrombosis following AstraZeneca COVID-19 Vaccination: Guidance Statement from the GTH. Hamostaseologie 2021. DOI: 10.1055/a-1469-7481.

