## Supplementary Appendix for "Anti-PF4 VITT antibodies are oligoclonal and variably inhibited by heparin"

David Murray, MD PhD

Division of Clinical Biochemistry & Immunology

Department of Laboratory Medicine & Pathology

200 1st Street SW

Mayo Clinic

Rochester, MN 55906

Anand Padmanabhan, MBBS PhD

Divisions of Hematopathology, Transfusion Medicine and Experimental Pathology

Department of Laboratory Medicine & Pathology

200 1st Street SW

Mayo Clinic

Rochester, MN 55906

### Table of Contents


| | Page |
|---|---|
| **VITT Patient Histories** | **4** |
| **Methods** | **8** |
| **Figures** | **10** |
| **References** | **15** |

***VITT patient clinical histories***

**Patient 1**'s clinical course has partially been presented previously[1]. Briefly, this patient was in the 50s (yrs) with no significant past medical history who presented with conjunctival congestion, retro-orbital pain, and diplopia ten days after ChAdOx1 nCoV-19 vaccination. Imaging revealed superior ophthalmic vein thrombosis (SOVT). Platelet count was $30 \times 10^9$/L (**Fig S1A**) and testing was consistent with ITP following a positive platelet suspension immunofluorescence test and positive monoclonal antibody-specific immobilization of platelet antigens (MAIPA) assay (data not shown). IgG antibodies against platelet factor 4 (PF4) were negative in lateral flow immunoassay testing (STIc Expert, Stago). Eight days after admission, the patient developed a transient, mild, right-sided hemiparesis, and aphasia. Magnetic resonance imaging (MRI) showed a new ischemic stroke in the left parietal lobe. The patient developed right-sided focal seizures adjudicated as related to the ischemic stroke which was treated successfully with levetiracetam and lacosamide. Approximately 10 days post-admission D-dimer measured was elevated at 17,024 ng/mL FEU. Patient was treated with unfractionated heparin and dexamethasone upon admission which was associated with an increase in platelet count during the treatment period. Upon platelet recovery, anticoagulation was switched to phenprocoumon with enoxaparin bridging and the patient was discharged on day 26 post-admission. Approximately 3 months after initial presentation, patient presented with symptoms consistent with focal seizures of the left hand. MRI revealed thrombosis in the anterior part of the superior sagittal sinus and draining veins on the left side, and in the left sigmoid sinus. D-dimer was normal, and international normalized ratio (INR) was in the therapeutic range (2 to 3). Cerebral venous sinus thrombosis (CVST) was not evident on the initial MRI at presentation. While precise time of development of CVST is unknown, one possibility includes the 2-week period after admission when platelets had not fully recovered during which anticoagulation with unfractionated heparin was measured as sub-therapeutic incidentally followed by dose adjustments.

**Patient 2**'s clinical history is partially presented elsewhere[2]. Briefly, this patient was in the 40s (yrs) with a history of asthma who presented with a one-week history of bilateral lower extremity pain that started eleven days after receiving the Ad26.COV2.S vaccine. At presentation, patient was noted to have a platelet count of 74 × $10^9$/L, fibrinogen activity of 254 mg/dL, D-dimer of 15,109 ng/mL FEU (**Fig S1B**). Venous duplex ultrasound of the lower extremities revealed bilateral occlusive and extensive deep venous thromboses (DVTs). A computerized tomography (CT) scan of the chest revealed multiple acute bilateral pulmonary emboli in the segmental and more proximal arteries. Magnetic resonance venography (MRV) and angiography (MRA) of the brain were unremarkable. A hypercoagulable work-up did not reveal any abnormalities. A presumptive diagnosis of VITT was made, and the patient was treated with 1 g/kg of intravenous immunoglobulins (IVIG) for 2 days, 1 mg/kg of prednisone daily, and intravenous argatroban infusion drip. His anti-PF4 enzyme-linked immunosorbent assay (ELISA) (LIFECODES PF4 IgG, Immucor) performed on a pre-treatment sample demonstrated a strongly positive result of 3.323 optical density (OD) units (reference interval < 0.399), consistent with a diagnosis of VITT. Serotonin release assay (SRA; 60% release with 0.1 U/mL unfractionated heparin, 3% release with 100 U/mL unfractionated heparin) and P-selectin expression assay (PEA; 76% expression with 30 mcg/mL PF4, 0% expression with 100 U/mL unfractionated heparin) were both positive. The patient's leg and chest pain resolved, his platelet count normalized on the third day of hospitalization, and patient was discharged on apixaban 10 mg twice daily. Over the subsequent month, the patient developed recurrent thrombocytopenia, with a platelet count that decreased from a peak of 205 × $10^9$/L to 107 × $10^9$/L. The patient maintained persistently strong positive anti-PF4 results (range: 2.4 – 2.8 OD units); however, repeat D-dimer levels, SRA, and PEA were all negative, which were interpreted to suggest that there was no further ongoing platelet activation. The patient's thrombocytopenia appeared to moderately correlate with prednisone dose adjustments but did not respond to repeat IVIG administration.

**Patient 3** was a patient in the 30s (yrs) who developed increasingly severe headaches with blurry vision, neck pain, stiffness nausea and vomiting approximately 2 weeks after receiving the Ad26.COV2.S vaccine. MRI demonstrated a large hematoma in the right posterior temporal lobe with surrounding edema and associated mass-effect with a right-to-left midline shift. Thrombi were also noted within the torcula, right transverse sinus, right sigmoid sinus, and upper internal jugular vein. At the time, the patient had a platelet count of $102 \times 10^9$/L (**Fig S1C**) with an elevated D-dimer of more than 40,000 ng/mL FEU and a normal fibrinogen activity (386mg/dL). The patient was started on IVIG (1 g/kg x 2 doses) and underwent chemical thrombolysis utilizing direct administration of TPA followed by a balloon thrombectomy. Bivalirudin was used as the anticoagulant during and after the procedure. The patient was also started on prednisone. The patient made a complete neurological recovery and was discharged 10 days later with a steroid taper and on apixaban.

**Patient 4** was a patient in the 40s (yrs) who received the Ad26.COV2.S vaccine. Two weeks later the patient experienced discomfort in his right lower extremity and a headache. A lower extremity ultrasound was negative for DVT, and CT of the head was unremarkable. The patient's platelet count was $25 \times 10^9$/L (**Fig S1D**). Two days later the patient had worsening left lower extremity LLE pain and swelling, as well as ongoing headaches. Imaging was repeated; the patient did not have a cerebral sinus thrombosis but had developed a DVT (left popliteal vein) and pulmonary embolism. D-dimer was elevated at 28,000 ng/mL FEU and fibrinogen was 200mg/dL. The patient was admitted and treated with prednisone, IVIG (1 g/kg x 2 doses) and bivalirudin. The patient was eventually discharged on apixaban and with a prednisone taper.

**Patient 5** was a patient in the 40s (yrs) who developed symptoms of headache and neck pain 2 weeks after receiving the Ad26.COV2.S vaccine, presented previously[3]. The patinet was found to have an occlusive cerebral sinus venous thrombosis (CSVT) involving the left transverse sigmoid sinus and internal jugular vein. There was no brain infarction. A CT angiogram of the chest demonstrated subsegmental filling defects in the right lung, involving the posterior basal segment and the superior segment of the right lower lobe. There were no other sites of thrombosis. On admission, the patient was thrombocytopenic with a platelet count of 20 000× $10^9$/L with an elevated D-dimer level of 27,150 ng/ml FEU and a fibrinogen of 149 mg/dL (**Fig S1E**). The patient was started on bivalirudin, daily prednisone (1 mg/kg) and intravenous immune globulin (IVIG) dosed at 1 g/kg/day for 2 days. The patient remained clinically stable with no additional signs or symptoms. By hospital day 6, the patient's platelets had risen to 115 000× $10^9$/L, and was transitioned to and discharged on rivaroxaban and a prednisone taper. On most recent follow up, 3 months later, the patient remains clinically stable with a platelet count of 283× $10^9$/L and a D dimer of <250 ng/ml FEU.

**Methods**

*Liquid Chromatography Electrospray Ionization Quadrupole time-of-flight mass spectrometry (LC-ESI-QTOF MS)*

The basic method was as described elsewhere[4,5]. Immunoglobulins (Igs) from patient sera or bead eluates were isolated using camelid-derived nanobodies directed against the constant domains of gamma heavy chain, kappa light chain and lambda light chain (Thermo Fisher Scientific). 10 µL of camelid nanobody beads were incubated with 20 µL of serum or 50 µL of PF4-Heparin Sepharose (or control) bead eluate diluted into 200 µL of buffered saline (PBS) for 45 minutes at ambient temperature. Subsequently, the supernatant was removed, and the beads were washed three times with 500 µL of water. Bound immunoglobulin light/heavy chains were eluted with 100 µL of 5% acetic acid and combined with 50 µL of 100 mM dithiothreitol (DTT) in 1M ammonium bicarbonate to disassociate Immunoglobulins into separated light chain and heavy chain components. An Eksigent Ekspert 200 microLC (Framingham, MA) liquid chromatography system was used to separate immunoglobulin chains prior to ionization and detection (which removes eluted PF4 prior to mass spectrometry). The mobile phases included an aqueous phase A (100% water + 1% formic acid) and an organic phase B (90% acetonitrile + 10% isopropanol + 0.1% formic acid). 5 µL of each camelid nanobody bead elution was injected per analysis onto a Poroshell 300SB-C3 column (1.0 mm X 75 mm) with a 5 µm particle size placed in a 60 ºC column heater. The gradient used has been described previously. The flow rate was 25 µL/minute. A SCIEX TripleTOF 5600 quadrupole time-of-flight (Q-TOF) mass spectrometer using electrospray ionization in positive ion mode was used for analysis. Data analysis was performed using Analyst TF v1.8.1 and PeakView ver. 2.2. Overexpressed Ig were inferred from the light chain +11 (m/z, mass to charge 2020 to m/z 2200) as described elsewhere[4,5]. The mass spectra of the multiply charged LC ions were deconvoluted to accurate molecular mass using the Bio Tool Kit ver. 2.2 plug-in software. The retention time of the monoclonal LC in each pre-treatment patient sample was tracked using PeakView. The instrument was calibrated every 5 samples using the automated calibrant delivery system (CDS). Mass measurement accuracy was estimated to be 15 ppm over the course of the analysis.

#### Supplementary Figure S1

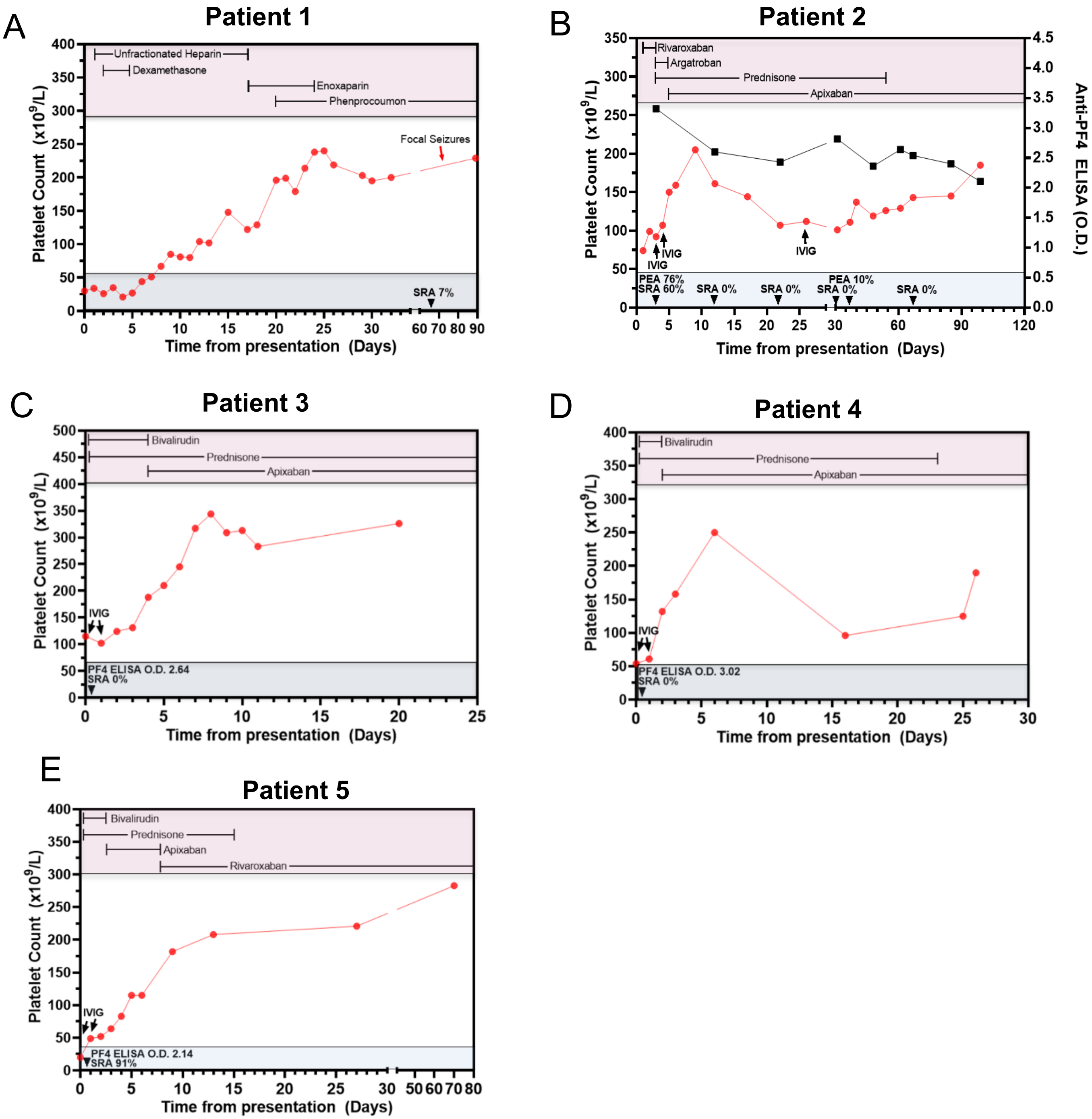


**Supplementary Figure S1. Clinical Course of VITT patients.** Left ordinate depicts platelet counts (red circles). Treatment interventions are displayed at the top of each panel (red box), and black arrows denote IVIG treatment (if given). For Patient 2, right ordinate denotes anti-PF4 ELISA results (black squares). Additional clinical results are presented at the bottom of each panel (gray box) with the time post-presentation denoted by black arrowheads. (**A-E**) Clinical course of Patients 1 through 5, respectively.

**Supplementary Figure S2**

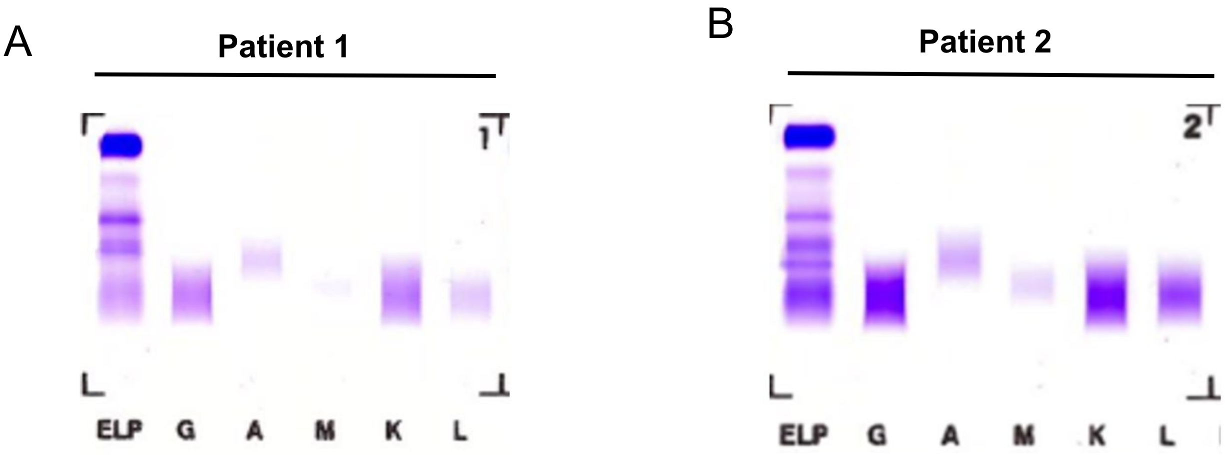


**Supplementary Figure S2. Immunofixation electrophoresis of native VITT samples does not demonstrate mono/oligoclonal bands**. Serum samples from Patient 1 **(A)** and Patient 2 (**B**) were run by electrophoresis with immunofixation to detect IgG, IgA, IgM, kappa, and lambda light chains following manufacturer instructions. In order, the gel lanes are presented as protein fixative (ELP), IgG (G), IgA (A), IgM (M), kappa light chain (K), and lambda light chain (L).

**Supplementary Figure S3**

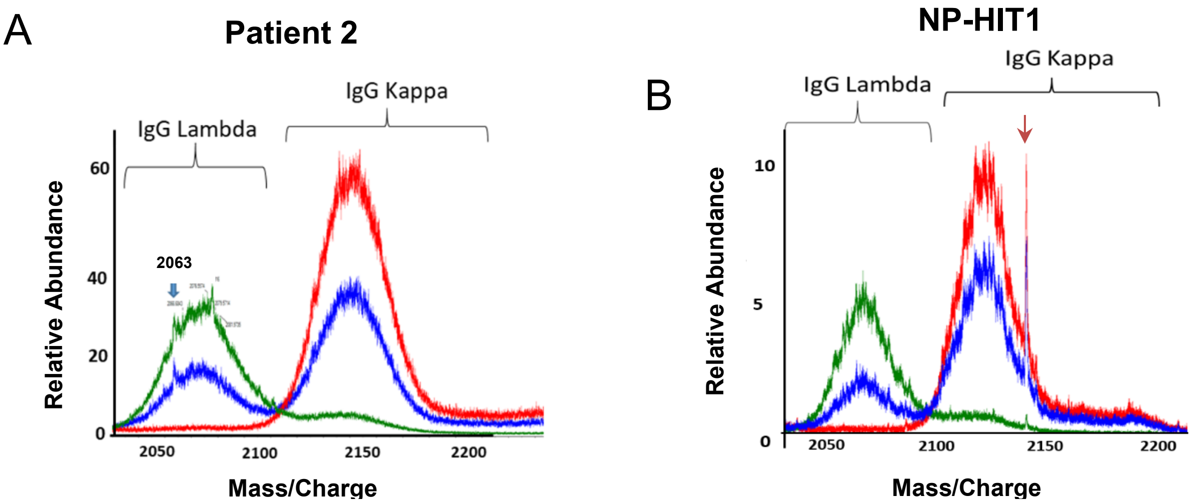


**Supplementary Figure S3. Mass Spectrometric evaluation of VITT and NP-HIT native sera.** Displayed are LC-ESI-QTOF MS +11 light chain distributions from native sera of VITT Patient 2 (**A**) and NP-HIT1 (with non-pathogenic anti-PF4 HIT antibodies) (**B**). In the spectra, green represents the distribution of all lambda containing immunoglobulins (Igs), red represents the distribution of all kappa containing Igs, and blue represents the light chain distribution of kappa and lambda light chains associated with an IgG heavy chain. The X-axis depicts mass/charge ratios and Y-axis depicts the relative abundance of the antibodies identified. The arrow in (**A**) depicts the monoclonal antibody identified in this patient after anti-PF4 isolation (**Fig 4B**), while the arrow in (**B**) shows an irrelevant monoclonal antibody in NP-HIT1 not pulled-down with PF4-heparin sepharose beads (**Fig 5E**).

Supplementary Figure S4

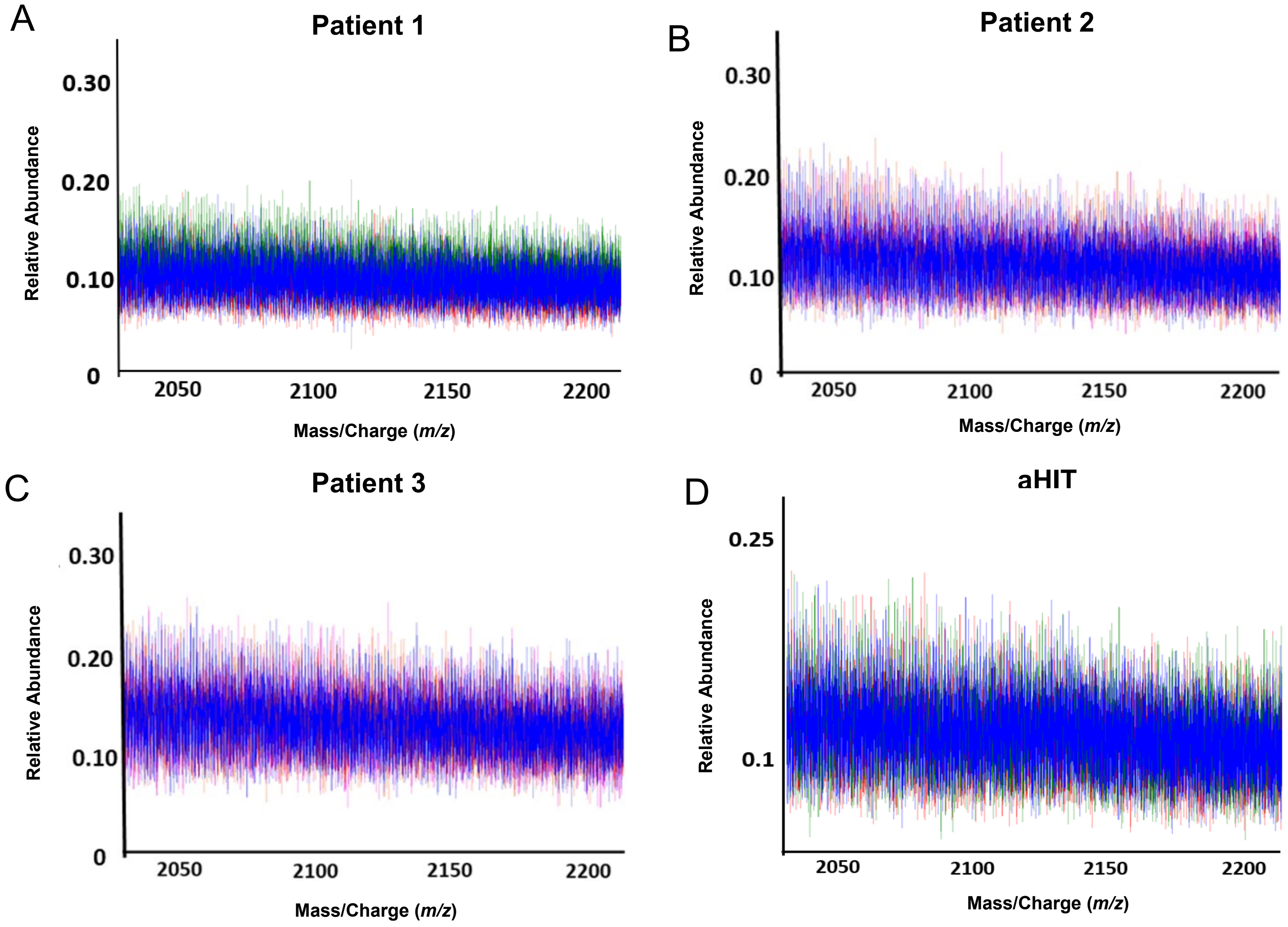


**Supplementary Figure 4. Elution from heparin-sepharose beads incubated with VITT and aHIT patient samples showed no Immunoglobulins in LC ESI QTOF MS.** Displayed are LC-ESI-QTOF MS +11 light chain distributions from anti-PF4 antibodies isolated patients with VITT (**A-C**) and Spontaneous HIT (aHIT) (**D**). In the spectra, green represents the distribution of all lambda containing immunoglobulins (Igs), red represents the distribution of all kappa containing Igs, and blue represents the light chain distribution of kappa and lambda light chains associated with an IgG heavy chain. The X-axis depicts mass/charge ratios and Y-axis depicts the relative abundance of the monoclonal/oligoclonal antibody identified. No Igs were seen.

**Supplementary Figure S5**

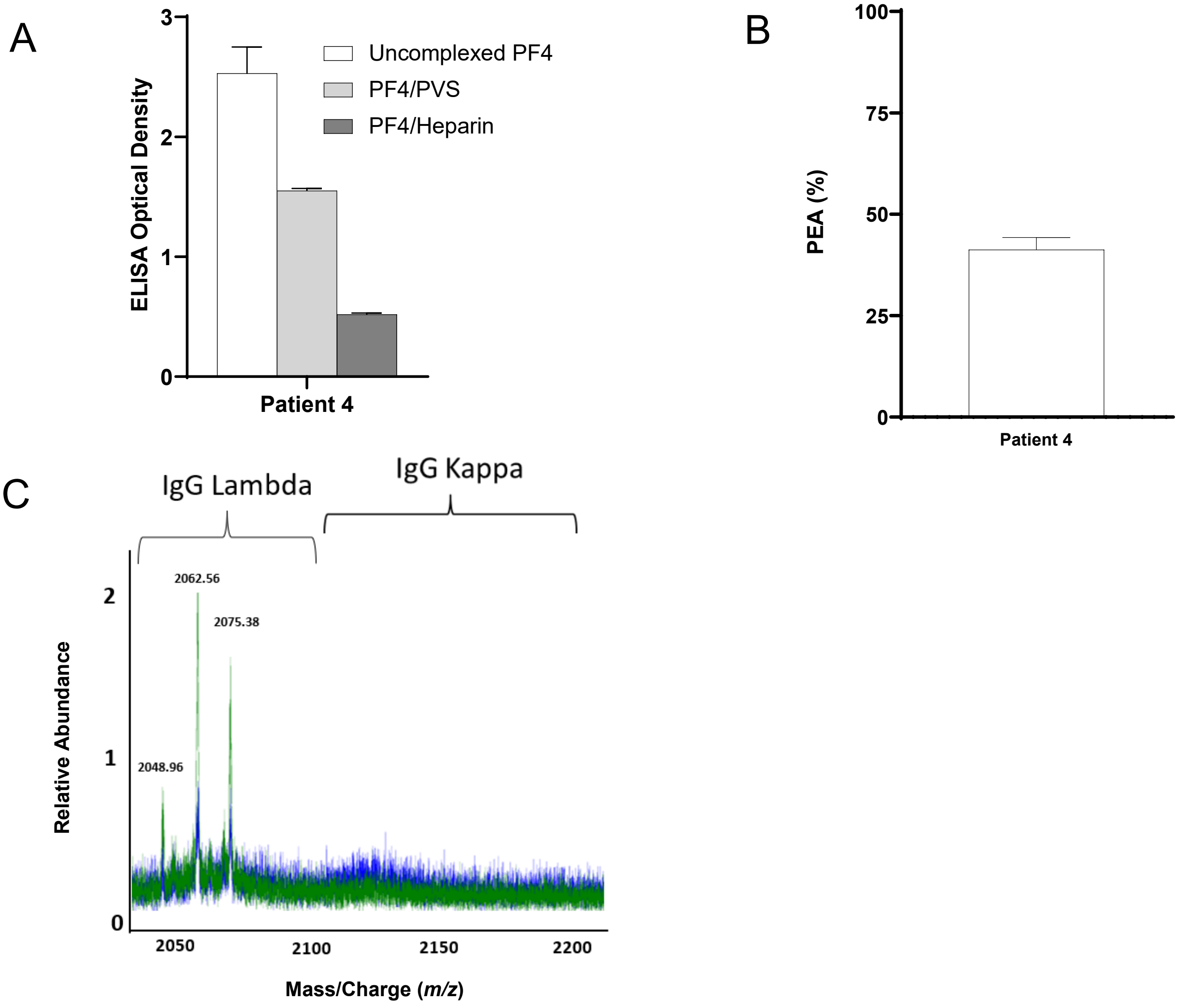


**Supplementary Figure 5. VITT antibodies are persistent.** Patient 4 sample was obtained and tested ~6 weeks after initial acute presentation. **(A)** Antibody binding to PF4 alone (“uncomplexed”; white), PF4/Polyvinylsulfonate (PVS; light gray), or PF4/Unfractionated heparin (dark gray) targets were evaluated by ELISA. **(B)** Activation of VITT patient antibodies in the PF4-dependent P-selectin Expression Assay (PEA) was examined. Mean and 1 SD are shown (n=3). **(C)** Displayed are LC-ESI-QTOF MS +11 light chain distributions from anti-PF4 antibodies isolated from this patient. In the spectra, green represents the distribution of all lambda containing immunoglobulins (Igs), red represents the distribution of all kappa containing Igs, and blue represents the light chain distribution of kappa and lambda light chains associated with an IgG heavy chain. The number listed above peaks indicate the mass/charge (m/z) ratio of the identified light chain. The X-axis shows mass/charge ratios and Y-axis depicts the relative abundance of the monoclonal/oligoclonal antibody identified.

**References**

1. Bayas A, Menacher M, Christ M, Behrens L, Rank A, Naumann M. Bilateral superior ophthalmic vein thrombosis, ischaemic stroke, and immune thrombocytopenia after ChAdOx1 nCoV-19 vaccination. Lancet 2021;397:e11.
2. Abou-Ismail MY, Moser KA, Smock KJ, Lim MY. Vaccine-induced thrombotic thrombocytopenia following Ad26.COV2.S vaccine in a man presenting as acute venous thromboembolism. Am J Hematol 2021.
3. George G, Friedman KD, Curtis BR, Lind SE. Successful treatment of thrombotic thrombocytopenia with cerebral sinus venous thrombosis following Ad26.COV2.S vaccination. Am J Hematol 2021.
4. Barnidge DR, Dasari S, Ramirez-Alvarado M, et al. Phenotyping polyclonal kappa and lambda light chain molecular mass distributions in patient serum using mass spectrometry. J Proteome Res 2014;13:5198-205.
5. Barnidge DR, Dasari S, Botz CM, et al. Using mass spectrometry to monitor monoclonal immunoglobulins in patients with a monoclonal gammopathy. J Proteome Res 2014;13:1419-27.
